# A Comparative Study of Acoustic Feature Extraction Using CSL and Web Prototype of LIS-N Application

**DOI:** 10.64898/2026.09.14.26362731

**Authors:** Shrramana Ganesh Sudhakar, Sabrina Musteric, Stephanie Watts, Mohamed Ebraheem, John Michael Templeton, Bridge2AI-Voice Consortium, Yaël Bensoussan

## Abstract

1.

**Purpose:** Conventional clinical acoustic analysis systems are confined to specialized clinical environments, which limits scalable and remote voice monitoring. This study aimed to validate LIS-N, a web-based prototype of a novel mobile application that uses the open-source Praat-Parselmouth library for acoustic feature extraction, against the Computerized Speech Lab (CSL), the clinical reference standard, to assess its potential for telehealth and longitudinal voice assessment before integration into clinical or research workflows.

**Method:** Twenty adult volunteers without voice complaints or diagnosed dysphonia completed three voice tasks across three consecutive days at a tertiary academic voice center in quiet isolated rooms. All participants completed the full protocol across three consecutive days. Voice tasks included Rainbow Passage reading, sustained vowel /i/, and maximum phonation time /a/. Audio was captured concurrently using CSL and the LIS-N prototype, which uses Praat-Parselmouth for acoustic feature extraction. To isolate the contributions of recording hardware and analysis software, four cross-system conditions were also examined. Agreement was assessed using Pearson correlation and Bland-Altman analyses. Session-to-session trajectory correlations were computed to evaluate temporal consistency across systems.

**Results:** Fundamental frequency and Pitch Mean demonstrated excellent agreement across all tasks and conditions (r > 0.98), with robustness to hardware and software differences. Pitch Minimum and Pitch Maximum showed strong agreement during sustained vowel tasks. Energy-based features showed moderate agreement, with variability driven primarily by microphone differences, and same-microphone conditions yielded substantially stronger energy agreement. Perturbation measures including jitter, shimmer and CPP showed consistently poor cross-platform agreement, consistent with prior literature.

**Conclusion:** This study provides proof of concept for the viability of open-source acoustic frameworks as accessible alternatives to proprietary clinical systems, while also identifying limitations. Both systems should be interpreted separately, as each remains reliable within its own framework. The LIS-N prototype demonstrated strong agreement for frequency-based voice measures and consistent longitudinal monitoring, supporting adoption of open-source acoustic tools in telehealth and remote voice monitoring.

## 2. Introduction

Voice is emerging as a clinically valuable signal, with acoustic measurements offering objective markers of health across medical domains [1]. Advances in voice science, telehealth, and machine learning (ML) have enabled disease detection, patient monitoring and extraction of quantifiable speech biomarkers [3,4]. Beyond their established links to specific disorders, acoustic features can now be analyzed at scale, enabling automated screening and longitudinal tracking of disease progression.

Conventional acoustic analysis systems used in voice clinics are resource-intensive and confined to controlled clinical environments, limiting routine or remote voice monitoring in real-world healthcare settings. Traditionally, these analyses are conducted in specialized clinical environments using proprietary platforms such as the CSL (Model 4500b, KayPENTAX, Montvale, NJ, USA). Although these types of systems are reliable, they require expensive equipment, trained personnel, and in-person visits. Because voice data collection and analysis are not fully integrated into a streamlined workflow, these systems can be time consuming and inefficient in clinical practice [5]. This limits the scalability of acoustic monitoring, particularly when frequent or remote assessments are needed.

Given the global expansion of telemedicine [6], there is an increasing need for accessible and clinically meaningful remote monitoring tools. Open-source packages like Praat-Parselmouth [2], a Python wrapper for the Praat software [7], offer robust acoustic feature extraction capabilities. However, few have been validated for clinical use or compared directly with gold-standard systems under real-world conditions [8].

To address this gap, our team developed LIS-N, a novel mobile application for acoustic analysis. For this comparative study, we used a browser-based LIS-N prototype that leverages Parselmouth. The tool allows users, including clinicians and patients, to record or upload audio and obtain speech metrics in real time. Unlike CSL, which operates in a closed hardware and software ecosystem, LIS-N emphasizes accessibility and workflow efficiency.

This study evaluates the reliability and clinical potential of LIS-N by comparing prototype acoustic outputs with CSL under concurrent recording conditions. The primary outcome is the cross-sectional agreement between LIS-N and CSL on key acoustic features extracted from the same vocal samples within a single session. Demonstrating sufficient agreement on this primary outcome would support LIS-N as a viable alternative to in-clinic based assessments.

The secondary outcomes are threefold. First, we assessed longitudinal consistency by comparing session to session trajectories between the systems, across three consecutive recording days. Although CSL and Praat based acoustic measures have been compared previously, prior work has not evaluated longitudinal consistency using concurrent recordings from both systems across consecutive days. Second, we examine the relative contributions of recording hardware versus analysis software to any observed disagreement through a cross-system analysis of four configurations (CSL microphone with CSL software, headset microphone with Praat-Parselmouth, CSL microphone with Praat-Parselmouth, and headset microphone with CSL software), in order to determine whether differences between systems are driven primarily by the microphone, the analysis pipeline, or both. Third, we evaluate which categories of acoustic features (frequency-based, energy-based, or perturbation/spectral) are most robust to these hardware and software differences, to inform feature selection and microphone standardization in remote acoustic assessment protocols.

## 3. Materials & Method

### 3.1. Data Collection

A total of twenty adult participants were recruited for this study from a convenience sample of medical students and research fellows with no voice complaints or diagnosed dysphonia. This study was deemed exempt and Not Human Research Subject by the University of South Florida Institutional Review Board. A healthy volunteer cohort was selected for this preliminary validation to establish baseline system agreement under relatively stable voice conditions before testing the platform in dysphonic populations. This was particularly important because acoustic measures in dysphonic voices can show substantial variability and may complicate initial validation. Each participant was instructed to complete three speech tasks repeated over three consecutive days. The tasks included reading a standardized passage known as the Rainbow Passage, producing a sustained vowel sound (/i/), and performing a maximum phonation time task with vowel sound (/a/). These tasks were chosen to capture both connected speech and sustained phonatory behavior, which are commonly used in clinical voice assessments.

All audio recordings were captured simultaneously using two systems for comparison (shown in Figure 1). All sessions were conducted in a quiet environment to minimize background noise and ensure consistency in audio quality. The first system was the CSL, a commercially available platform widely used in clinical voice analysis. The second is a web-based prototype of LIS-N app that uses the Praat-Parselmouth library.

**Figure 1.**
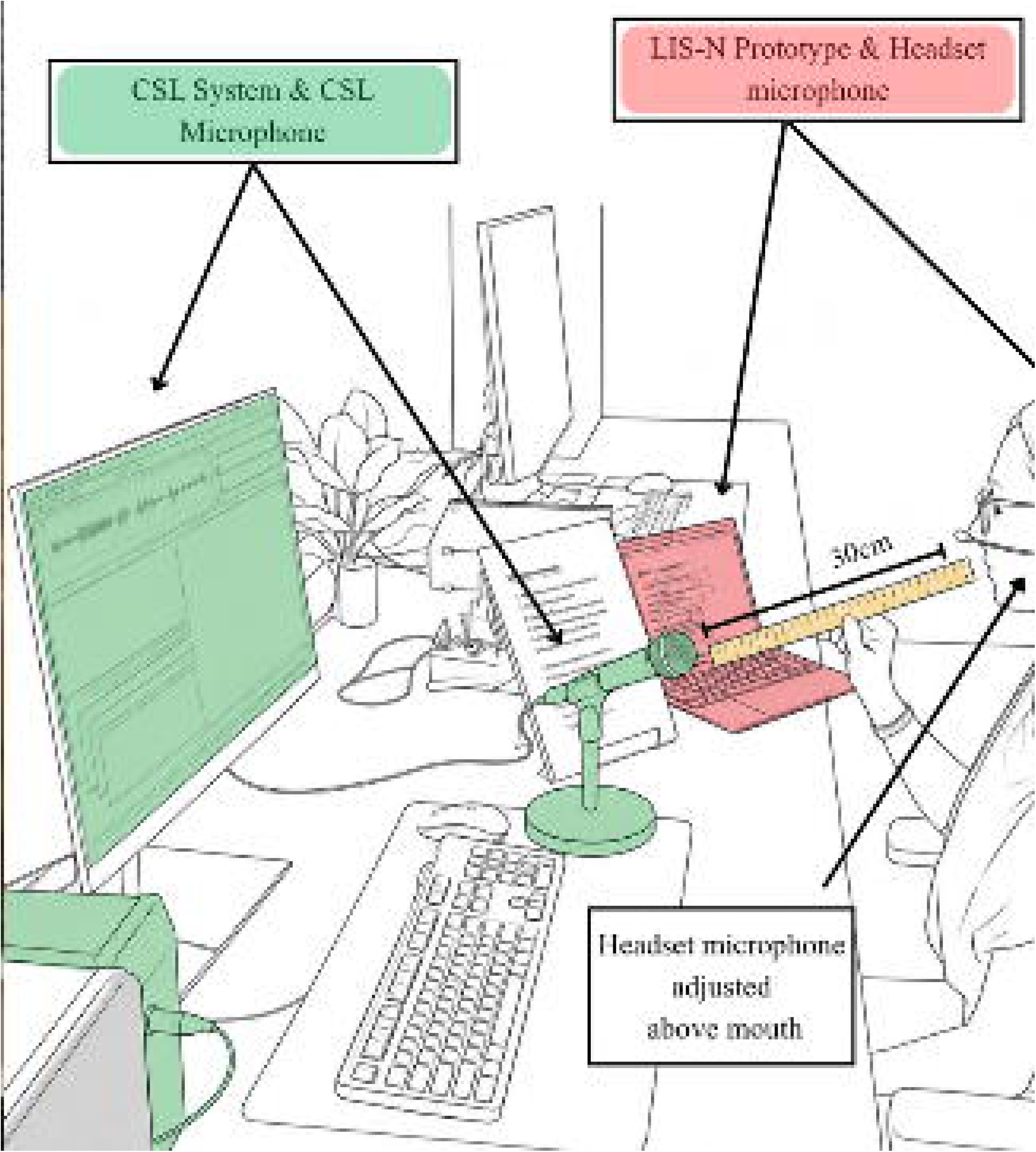
Concurrent Audio Recording Protocol Using the Computerized Speech Lab and the LIS-N Prototype.

Recordings for the CSL system were made using the standard microphone provided with the system, positioned approximately twelve inches in front of the participant’s mouth, as per conventional clinical guidelines. For LIS-N prototype, a headset microphone was used instead [9]. This microphone was positioned just above the mouth to avoid direct airflow, which can introduce distortion, particularly during the production of plosive sounds such as "p".

Before each recording, participants were asked to perform a short vocalization of the sequence "pa-pa-pa" to identify and reduce the presence of plosive artifacts in the signal. The angle of the headset was adjusted as needed based on this check [10].

### 3.2. LIS-N Prototype’s Web Interface and Acoustic Analysis Pipeline

The platform allows participants or clinicians to record, upload pre-recorded audio, interactively trim recordings and extract acoustic features within a unified interface.

While modern web browsers allow direct microphone access, preliminary testing revealed that using Streamlit [17] did not give direct control over the microphone and few browsers apply automatic gain control (AGC) to incoming audio. This AGC introduces inconsistencies in signal amplitude, potentially affecting the reliability of extracted acoustic features. To address this, a dedicated local recording application was developed (shown in Figure 2). This standalone tool enables participants to record high-quality audio without browser-based gain adjustments. The application also collects metadata such as participant identifier and date and saves the audio locally in uncompressed WAV format.

**Figure 2.**
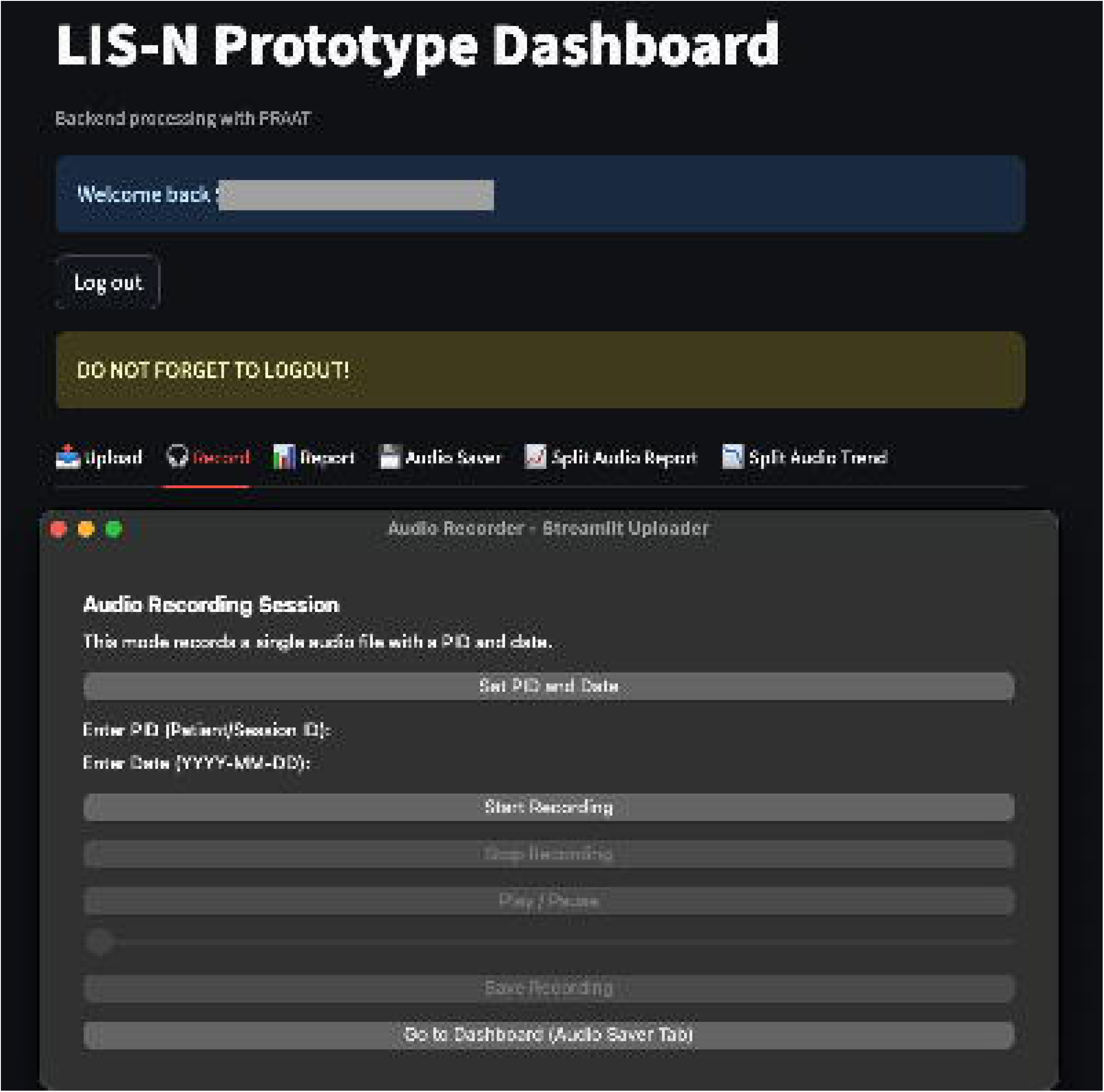
Offline Local Recording Application and LIS-N Prototype Web Dashboard Interface.

After the recording is completed, the application automatically redirects the user to the Streamlit-based web dashboard. Once logged in, the user can upload the locally recorded audio for further processing and the interface includes a trimming tool that allows users to isolate relevant portions (shown in Figure 3) of each recording and label them according to the associated speech task (e.g., Rainbow Passage, sustained vowel or maximum phonation time).

**Figure 3.**
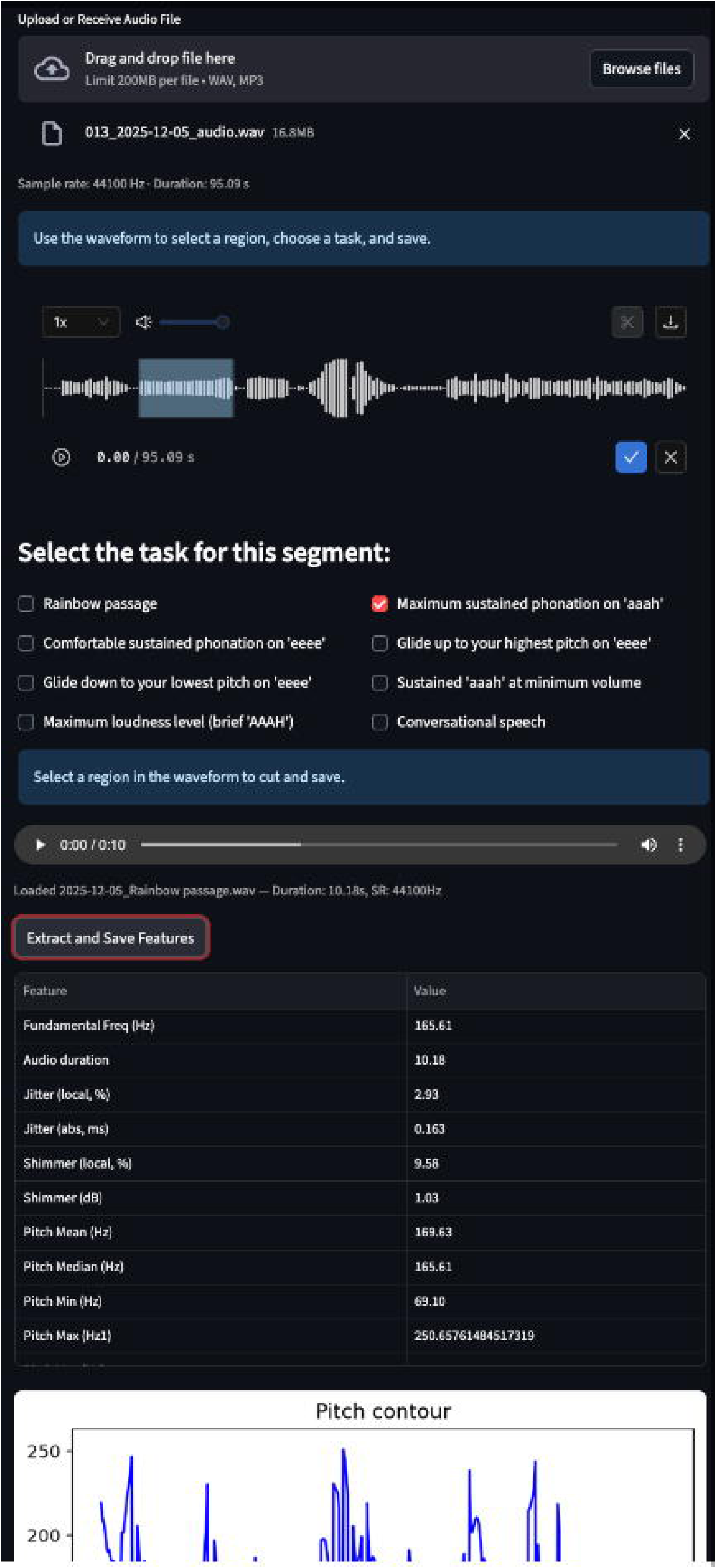
LIS-N Prototype Audio Trimming and Acoustic Feature Extraction Interface.

Acoustic analysis is performed within the same platform using Parselmouth. For this study, we used development version praat_parselmouth-0.5.0.dev0-cp313-cp313 manylinux_2_17_x86_64.manylinux2014_x86_64.whl, which includes the latest updates to pitch extraction algorithms and improved integration with Praat’s core functions. The dashboard also supports downloading processed results or storing them securely in a cloud repository.

This modular dashboard pipeline allows for efficient remote data collection, preprocessing and feature extraction, making it suitable for both research and potential clinical deployment.

### 3.3. Audio capture and Recording settings

In the CSL system, task segments are selected manually after recording. The technician has to manually adjust gain before recording, locate the relevant portions using the software interface, extract features through different CSL programs (MDVP, ADSV and Real-Time Pitch) and save the results manually. In the LIS-N prototype, while it supports recording and analyzing each task individually, this feature was not used in order to maintain protocol consistency. Instead, the complete recording was uploaded to the web interface, where it was trimmed into separate segments for each task to mirror the protocol with CSL. These segments were analyzed automatically and the extracted results were saved with the appropriate task labels.

All recordings were saved in standard uncompressed WAV format with a sampling rate of 44100 hertz. Each file was anonymized and labeled with a unique identifier to ensure objectivity during analysis. Since both the methods involved trimming the audio segments manually, there is some discrepancy in the selected segments.

For the LIS-N prototype, acoustic features were extracted from Parselmouth. Pitch was extracted using the filtered autocorrelation method with a pitch floor of 30 Hz, a ceiling of 600 Hz. All other settings followed the default configuration in the version used, which was Praat-Parselmouth version 0.5.0.dev0. CSL recordings were made using the default settings provided by the software, which were kept consistent across all sessions. Both these systems are capable of extracting a wide array of features and for this trial, the following features were extracted from each recording segment: Fundamental Freq (Hz), Audio duration, Jitter (local, %), Jitter (abs, ms), Shimmer (local, %), Shimmer (dB), Pitch Mean (Hz), Pitch Min (Hz), Pitch Max (Hz), Pitch Range (Hz), Energy Mean (dB), Energy Min (dB), Energy Max (dB), Energy Range (dB), CPP (dB).

### 3.4. Statistical analysis

To evaluate agreement between the two systems, acoustic feature values for each participant were averaged across the three recording sessions. Pearson correlation coefficients were then computed between the CSL and Parselmouth-based systems for each extracted feature to assess the strength of linear association. Bland-Altman analyses were additionally performed to quantify systematic bias and limits of agreement between the two systems.

To isolate the respective contributions of recording hardware and analysis software to any observed disagreement, a cross-system analysis was conducted across four conditions: CSL microphone audio analyzed with CSL software (CSL), LIS-N prototype headset microphone audio analyzed with Praat-Parselmouth (PRAAT), CSL microphone audio analyzed with Praat-Parselmouth (M_PRAAT), and LIS-N prototype headset microphone audio analyzed with CSL software (H_CSL). Pearson correlations and Bland-Altman analyses were computed for all pairwise comparisons among these conditions, following the same averaging procedure described above.

Finally, to assess temporal consistency, session-to-session trajectory correlations were computed for each participant and acoustic feature. Specifically, the three-session value trajectory of one system was correlated with the corresponding trajectory of the other system using Pearson correlation, capturing whether both systems tracked the same directional pattern of change across sessions rather than simply agreeing on absolute values. Individual participant correlation coefficients were aggregated using Fisher z-transformation to derive a mean r for each feature and system comparison. This trajectory analysis was applied across all four system comparison conditions to further characterize the effect of hardware and software differences on longitudinal measurement consistency.

## 4. Results

Twenty adult participants were enrolled (16 women [80%]; 4 men [20%]; mean [SD] age, 33.7 [12.5] years; range, 20–60 years). All participants were trainees, medical students, or research fellows, with no voice complaints or diagnosed dysphonia.

Agreement between the CSL and the LIS-N prototype varied across acoustic parameters and recording configurations. For the primary outcome of cross-sectional agreement, frequency-based measures showed excellent agreement between systems, energy-based measures showed moderate agreement that varied with microphone configuration, and perturbation and cepstral measures showed poor agreement across all conditions (Table 1).

**Table 1.** Pearson Correlation Coefficients for Acoustic Feature Agreement Between the LIS-N Prototype and Computerized Speech Lab Across Four Cross-System Configurations and Three Voice Tasks.

| Pearson Correlations — PRAAT vs CSL |  |  |  |  |  |  |
| --- | --- | --- | --- | --- | --- | --- |
| Measure | Rainbow Passage |  | MPT |  | Sustained Vowel |  |
|  | *r* | *p* | *r* | *p* | *r* | *p* |
| *Frequency-Based* |  |  |  |  |  |  |
| Fundamental Freq (Hz) | **0.987** | <.001<sup>c</sup> | **0.998** | <.001<sup>c</sup> | **0.995** | <.001<sup>c</sup> |
| Pitch Mean (Hz) | **0.986** | <.001<sup>c</sup> | **0.993** | <.001<sup>c</sup> | **0.995** | <.001<sup>c</sup> |
| Pitch Min (Hz) | 0.44 | 0.052 | **0.902** | <.001<sup>c</sup> | **0.841** | <.001<sup>c</sup> |
| Pitch Max (Hz) | 0.554 | 0.011<sup>a</sup> | 0.16 | 0.500 | **0.921** | <.001<sup>c</sup> |
| Pitch Range (Hz) | 0.357 | 0.122 | 0.311 | 0.182 | 0.318 | 0.172 |
| *Energy-Based* |  |  |  |  |  |  |
| Energy Mean (dB) | 0.075 | 0.752 | 0.672 | 0.001<sup>b</sup> | 0.691 | <.001<sup>c</sup> |
| Energy Max (dB) | 0.433 | 0.057 | 0.695 | <.001<sup>c</sup> | 0.59 | 0.006<sup>b</sup> |
| Energy Min (dB) | 0.204 | 0.387 | 0.072 | 0.763 | 0.476 | 0.034<sup>a</sup> |
| Energy Range (dB) | 0.299 | 0.201 | 0.524 | 0.018<sup>a</sup> | 0.603 | 0.005<sup>b</sup> |
| *Perturbation & Spectral* |  |  |  |  |  |  |
| Jitter (local, %) | 0.38 | 0.099 | 0.427 | 0.061 | -0.239 | 0.309 |
| Jitter (abs, ms) | 0.494 | 0.027<sup>a</sup> | 0.693 | <.001<sup>c</sup> | -0.356 | 0.124 |
| Shimmer (local, %) | 0.646 | 0.002<sup>b</sup> | 0.39 | 0.089 | -0.174 | 0.464 |
| Shimmer (dB) | 0.59 | 0.006<sup>b</sup> | 0.377 | 0.101 | -0.172 | 0.467 |
| CPP (dB) | 0.066 | 0.781 | 0.022 | 0.926 | 0.243 | 0.303 |
| Pearson Correlations — M_PRAAT vs CSL |  |  |  |  |  |  |
| Measure | Rainbow Passage |  | MPT |  | Sustained Vowel |  |
|  | *r* | *p* | *r* | *p* | *r* | *p* |
| *Frequency-Based* |  |  |  |  |  |  |
| Fundamental Freq (Hz) | **0.994** | <.001<sup>c</sup> | **0.998** | <.001<sup>c</sup> | **0.987** | <.001<sup>c</sup> |
| Pitch Mean (Hz) | **0.989** | <.001<sup>c</sup> | **0.993** | <.001<sup>c</sup> | **0.983** | <.001<sup>c</sup> |
| Pitch Min (Hz) | 0.63 | 0.003<sup>b</sup> | **0.778** | <.001<sup>c</sup> | 0.667 | 0.001<sup>b</sup> |
| Pitch Max (Hz) | 0.577 | 0.008<sup>b</sup> | 0.198 | 0.402 | **0.871** | <.001<sup>c</sup> |
| Pitch Range (Hz) | 0.436 | 0.055 | 0.254 | 0.280 | -0.041 | 0.865 |
| *Energy-Based* |  |  |  |  |  |  |
| Energy Mean (dB) | **0.935** | <.001<sup>c</sup> | **0.983** | <.001<sup>c</sup> | **0.952** | <.001<sup>c</sup> |
| Energy Max (dB) | **0.977** | <.001<sup>c</sup> | **0.998** | <.001<sup>c</sup> | **0.974** | <.001<sup>c</sup> |
| Energy Min (dB) | 0.607 | 0.004<sup>b</sup> | 0.47 | 0.037<sup>a</sup> | 0.583 | 0.007<sup>b</sup> |

| Pearson Correlations — M_PRAAT vs CSL |  |  |  |  |  |  |
| --- | --- | --- | --- | --- | --- | --- |
| Measure | Rainbow Passage |  | MPT |  | Sustained Vowel |  |
|  | *r* | *p* | *r* | *p* | *r* | *p* |
| *Energy-based* |  |  |  |  |  |  |
| Energy Range (dB) | 0.63 | 0.003<sup>b</sup> | **0.741** | <.001<sup>c</sup> | **0.719** | <.001<sup>c</sup> |
| *Perturbation & Spectral* |  |  |  |  |  |  |
| Jitter (local, %) | 0.472 | 0.036<sup>a</sup> | 0.42 | 0.066 | -0.121 | 0.610 |
| Jitter (abs, ms) | 0.491 | 0.028<sup>a</sup> | **0.716** | <.001<sup>c</sup> | -0.192 | 0.418 |
| Shimmer (local, %) | 0.04 | 0.869 | 0.114 | 0.633 | -0.088 | 0.712 |
| Shimmer (dB) | -0.093 | 0.695 | 0.127 | 0.593 | -0.12 | 0.615 |
| CPP (dB) | -0.428 | 0.060 | 0.186 | 0.431 | 0.299 | 0.201 |
| Pearson Correlations — PRAAT vs H_CSL |  |  |  |  |  |  |
| Measure | Rainbow Passage |  | MPT |  | Sustained Vowel |  |
|  | *r* | *p* | *r* | *p* | *r* | *p* |
| *Frequency-Based* |  |  |  |  |  |  |
| Fundamental Freq (Hz) | **0.992** | <.001<sup>c</sup> | **1** | <.001<sup>c</sup> | **0.995** | <.001<sup>c</sup> |
| Pitch Mean (Hz) | **0.995** | <.001<sup>c</sup> | **1** | <.001<sup>c</sup> | **0.995** | <.001<sup>c</sup> |
| Pitch Min (Hz) | 0.607 | 0.005<sup>b</sup> | **0.964** | <.001<sup>c</sup> | **0.834** | <.001<sup>c</sup> |
| Pitch Max (Hz) | 0.663 | 0.001<sup>b</sup> | 0.669 | 0.001<sup>b</sup> | **0.964** | <.001<sup>c</sup> |
| Pitch Range (Hz) | 0.388 | 0.091 | 0.66 | 0.002<sup>b</sup> | 0.321 | 0.168 |
| *Energy-Based* |  |  |  |  |  |  |
| Energy Mean (dB) | **0.947** | <.001<sup>c</sup> | **0.966** | <.001<sup>c</sup> | **0.771** | <.001<sup>c</sup> |
| Energy Max (dB) | 0.461 | 0.041<sup>a</sup> | **0.918** | <.001<sup>c</sup> | **0.923** | <.001<sup>c</sup> |
| Energy Min (dB) | 0.407 | 0.075 | 0.561 | 0.010<sup>a</sup> | 0.193 | 0.414 |
| Energy Range (dB) | -0.014 | 0.954 | 0.549 | 0.012<sup>a</sup> | 0.385 | 0.093 |
| *Perturbation & Spectral* |  |  |  |  |  |  |
| Jitter (local, %) | 0.289 | 0.216 | 0.492 | 0.027<sup>a</sup> | -0.064 | 0.789 |
| Jitter (abs, ms) | 0.136 | 0.569 | 0.53 | 0.016<sup>a</sup> | -0.092 | 0.700 |
| Shimmer (local, %) | 0.608 | 0.004<sup>b</sup> | 0.368 | 0.111 | -0.102 | 0.669 |
| Shimmer (dB) | 0.527 | 0.017<sup>a</sup> | 0.345 | 0.137 | -0.101 | 0.670 |
| CPP (dB) | -0.479 | 0.033<sup>a</sup> | -0.018 | 0.940 | 0.322 | 0.166 |

| Pearson Correlations — M_PRAAT vs H_CSL |  |  |  |  |  |  |
| --- | --- | --- | --- | --- | --- | --- |
| Measure | Rainbow Passage |  | MPT |  | Sustained Vowel |  |
|  | *r* | *p* | *r* | *p* | *r* | *p* |
| *Frequency-Based* |  |  |  |  |  |  |
| Fundamental Freq (Hz) | **0.996** | <.001<sup>c</sup> | **1** | <.001<sup>c</sup> | **0.987** | <.001<sup>c</sup> |
| Pitch Mean (Hz) | **0.993** | <.001<sup>c</sup> | **1** | <.001<sup>c</sup> | **0.983** | <.001<sup>c</sup> |
| Pitch Min (Hz) | 0.508 | 0.022<sup>a</sup> | **0.864** | <.001<sup>c</sup> | 0.585 | 0.007<sup>b</sup> |
| Pitch Max (Hz) | **0.781** | <.001<sup>c</sup> | 0.638 | 0.002<sup>b</sup> | **0.986** | <.001<sup>c</sup> |
| Pitch Range (Hz) | 0.607 | 0.005<sup>b</sup> | 0.412 | 0.071 | -0.06 | 0.803 |
| *Energy-Based* |  |  |  |  |  |  |
| Energy Mean (dB) | 0.263 | 0.263 | 0.63 | 0.003<sup>b</sup> | 0.588 | 0.006<sup>b</sup> |
| Energy Max (dB) | 0.227 | 0.335 | 0.538 | 0.014<sup>a</sup> | 0.429 | 0.059 |
| Energy Min (dB) | -0.144 | 0.545 | 0.221 | 0.349 | 0.236 | 0.317 |
| Energy Range (dB) | -0.32 | 0.169 | 0.44 | 0.052 | 0.439 | 0.053 |
| *Perturbation & Spectral* |  |  |  |  |  |  |
| Jitter (local, %) | 0.366 | 0.112 | 0.434 | 0.056 | -0.128 | 0.590 |
| Jitter (abs, ms) | 0.117 | 0.623 | 0.527 | 0.017<sup>a</sup> | -0.098 | 0.682 |
| Shimmer (local, %) | -0.019 | 0.937 | 0.19 | 0.422 | -0.15 | 0.529 |
| Shimmer (dB) | -0.14 | 0.556 | 0.26 | 0.268 | -0.16 | 0.500 |
| CPP (dB) | -0.626 | 0.003<sup>b</sup> | 0.235 | 0.319 | 0.314 | 0.177 |

**Abbreviations:** CPP, cepstral peak prominence; CSL, Computerized Speech Lab system (CSL microphone analyzed with CSL software); F0, fundamental frequency; H\_CSL, LIS-N headset microphone audio analyzed with CSL software; M\_PRAAT, CSL microphone audio analyzed with Praat-Parselmouth; MPT, maximum phonation time; PRAAT, LIS-N headset microphone audio analyzed with Praat-Parselmouth.
**Footnote:** Values represent Pearson correlation coefficients (*r*) computed from participant-level means averaged across three recording sessions (*n* = 20). □ *P* < .05 □ *P* < .01 □ *P* < .001

For the secondary outcomes, session-to-session trajectory correlations broadly paralleled the cross-sectional pattern, with frequency measures tracking consistently across systems and perturbation measures showing poor or inverse longitudinal agreement (Table 2). The four-condition cross-system analysis further indicated that energy measures tracked consistently only when the microphone was held constant, whereas perturbation disagreement persisted regardless of hardware or software configuration.

**Table 2.** Mean Session-to-Session Trajectory Correlations Between Systems Across Four Cross-System Configurations and Three Voice Tasks.

|  | Session Trajectory Correlations:<br>CSL vs PRAAT |  |  | CSL vs M_PRAAT |  |  |
| --- | --- | --- | --- | --- | --- | --- |
| Measure | Rainbow<br>Passage | MPT | Sustained<br>Vowel | Rainbow<br>Passage | MPT | Sustained<br>Vowel |
| *Frequency-Based* |  |  |  |  |  |  |
| Fundamental Freq (Hz) | **0.854** | **0.998** | **1.000** | **0.800** | **0.998** | **0.999** |
| Pitch Mean (Hz) | **0.842** | **0.999** | **0.999** | **0.938** | **0.997** | **0.997** |
| Pitch Min (Hz) | 0.180 | **0.981** | **0.884** | -0.366 | **0.921** | **0.902** |
| Pitch Max (Hz) | 0.205 | **0.878** | **0.773** | -0.095 | **0.976** | **0.948** |
| Pitch Range (Hz) | 0.102 | 0.402 | 0.461 | -0.305 | **0.865** | 0.542 |
| *Energy-Based* |  |  |  |  |  |  |
| Energy Mean (dB) | 0.290 | 0.605 | 0.171 | **0.925** | **0.996** | **0.952** |
| Energy Max (dB) | 0.205 | **0.716** | 0.548 | **0.988** | **0.999** | **0.998** |
| Energy Min (dB) | -0.296 | -0.115 | 0.099 | 0.093 | 0.354 | **0.791** |
| Energy Range (dB) | -0.208 | 0.362 | 0.392 | **0.867** | 0.629 | **0.730** |
| *Perturbation & Spectral* |  |  |  |  |  |  |
| Jitter (local, %) | 0.024 | -0.035 | -0.053 | 0.110 | -0.114 | 0.220 |
| Jitter (abs, ms) | -0.095 | 0.240 | 0.100 | -0.241 | 0.116 | 0.494 |
| Shimmer (local, %) | -0.401 | 0.039 | -0.090 | -0.246 | **0.796** | **0.888** |
| Shimmer (dB) | 0.075 | 0.043 | 0.062 | -0.325 | **0.737** | **0.854** |
| CPP (dB) | 0.051 | **-0.738** | -0.523 | -0.121 | -0.253 | -0.504 |
|  | Session Trajectory Correlations:<br>PRAAT vs H_CSL |  |  | M_PRAAT vs H_CSL |  |  |
| Measure | Rainbow<br>Passage | MPT | Sustained<br>Vowel | Rainbow<br>Passage | MPT | Sustained<br>Vowel |
| *Frequency-Based* |  |  |  |  |  |  |
| Fundamental Freq (Hz) | **0.870** | **0.999** | **1.000** | **0.853** | **0.999** | **0.999** |
| Pitch Mean (Hz) | **0.777** | **1.000** | **1.000** | **0.814** | **0.999** | **0.999** |
| Pitch Min (Hz) | 0.232 | **0.988** | **0.986** | 0.122 | **0.952** | **0.936** |
| Pitch Max (Hz) | 0.523 | **0.793** | **0.832** | 0.112 | **0.926** | **0.947** |
| Pitch Range (Hz) | 0.412 | 0.336 | 0.376 | 0.177 | 0.621 | **0.733** |

|  | Session Trajectory Correlations:<br>PRAAT vs H_CSL |  |  | M_PRAAT vs H_CSL |  |  |
| --- | --- | --- | --- | --- | --- | --- |
| Measure | Rainbow<br>Passage | MPT | Sustained<br>Vowel | Rainbow<br>Passage | MPT | Sustained<br>Vowel |
| *Energy-Based* |  |  |  |  |  |  |
| Energy Mean (dB) | **0.844** | **0.970** | **0.890** | -0.159 | 0.333 | 0.225 |
| Energy Max (dB) | 0.603 | **0.910** | **0.987** | 0.042 | 0.578 | 0.266 |
| Energy Min (dB) | 0.071 | 0.228 | -0.002 | -0.352 | 0.145 | -0.614 |
| Energy Range (dB) | -0.264 | -0.045 | 0.512 | 0.254 | **0.744** | -0.032 |
| *Perturbation & Spectral* |  |  |  |  |  |  |
| Jitter (local, %) | -0.415 | -0.096 | 0.314 | 0.104 | -0.427 | 0.411 |
| Jitter (abs, ms) | -0.191 | 0.116 | 0.455 | 0.158 | 0.384 | 0.472 |
| Shimmer (local, %) | 0.692 | 0.183 | 0.684 | -0.197 | -0.504 | -0.247 |
| Shimmer (dB) | 0.444 | 0.081 | **0.819** | 0.012 | -0.151 | -0.213 |
| CPP (dB) | 0.480 | -0.548 | -0.478 | 0.308 | -0.171 | -0.402 |

**Abbreviations:** CPP, cepstral peak prominence; CSL, Computerized Speech Lab system (CSL microphone analyzed with CSL software); F0, fundamental frequency; H\_CSL, LIS-N headset microphone audio analyzed with CSL software; M\_PRAAT, CSL microphone audio analyzed with Praat-Parselmouth; MPT, maximum phonation time; PRAAT, LIS-N headset microphone audio analyzed with Praat-Parselmouth.
**Footnote:** Values represent mean within-participant Pearson correlation coefficients between the three-session trajectories of paired systems, aggregated across participants ( $n = 20$ ) using Fisher z-transformation. Higher values indicate greater concordance in directional change across recording sessions between the two systems compared. Bold values indicate $|r| \geq 0.70$ .

### 4.1. Frequency-Based Features

#### System Agreement

Fundamental frequency (F0) and Pitch Mean demonstrated excellent agreement between the two systems across all three tasks, with Pearson correlations exceeding r = 0.98 across all tasks and all four cross-system configurations (Table 1). This indicates that these measures are robust to both hardware and software differences.

In contrast, extreme pitch values (Pitch Minimum, Pitch Maximum and Pitch Range) showed task-dependent agreement. Agreement was substantially stronger during sustained phonation tasks than during connected speech. Notably, Pitch Minimum and Pitch Maximum dropped to r = 0.44 and r = 0.55 respectively during the Rainbow Passage in the standard comparison, while reaching r = 0.90 and r = 0.92 during MPT and sustained vowel. Pitch Range showed poor agreement across all tasks and conditions. This pattern was consistent across all four cross-system configurations (Table 1).

#### Temporal Consistency Across Sessions

Fundamental frequency and Pitch Mean demonstrated near-ceiling trajectory matching across all tasks and all system comparisons, with mean r values ranging from r = 0.78 to 1.00 (Table 2). Pitch Minimum and Pitch Maximum showed strong trajectory matching during MPT and sustained vowel across most conditions, though agreement weakened considerably for the Rainbow Passage, with some negative values observed in cross-system comparisons. Pitch Range showed poor trajectory consistency across all tasks and conditions.

### 4.2. Energy-Based Features

#### System Agreement

Energy features showed moderate agreement in the standard “PRAAT vs. CSL” comparison, with the notably low correlation of r = 0.08 for Energy Mean during the Rainbow Passage standing out as an unexpected finding (Table 1). Agreement improved substantially when the same microphone was used across both software platforms, with Energy Mean and Energy Maximum reaching r = 0.95 to 1.00 for MPT and sustained vowel in same-microphone conditions. In contrast, comparisons involving different microphones showed consistently weaker energy agreement, confirming that recording hardware is a primary driver of energy measurement variability. Notably, energy values were systematically higher when headset-recorded audio was analyzed with CSL software, while CSL microphone audio analyzed with Praat-Parselmouth yielded lower values than the same audio analyzed with CSL.

#### Temporal Consistency Across Sessions

Energy trajectory matching was strongly influenced by microphone consistency. When the same microphone was used across both software platforms, trajectory matching was strong across most tasks (Table 2). When microphones differed, trajectory agreement was substantially weaker and highly variable, with several near-zero and negative values observed.

### 4.3. Perturbation and Spectral Features

#### System Agreement

Jitter (local and absolute), Shimmer (local and dB), and CPP showed consistently poor agreement across all tasks and all four cross-system configurations (Table 1). Most notably, sustained vowel perturbation measures exhibited negative correlations in the standard comparison (e.g., Jitter absolute r = −0.36, Shimmer local r = −0.17), indicating an inverse relationship between the two system’s measurements. CPP showed near-zero correlations across all tasks and conditions.

#### Temporal Consistency Across Sessions

Perturbation measures and CPP showed poor or inverse trajectory matching across all tasks and all system comparisons, with mean r values frequently near zero or negative (Table 2). Most strikingly, CPP showed negative trajectory correlations across multiple conditions, reaching as low as r = −0.74 for MPT in the standard comparison. This pattern was consistent across all four cross-system conditions.

## 5. Discussion

The findings from this comparison provide early evidence that open-source acoustic analysis tools can perform comparably to established clinical systems for selected acoustic measures.

Agreement between the LIS-N prototype and CSL was feature dependent. Frequency based measures showed the strongest cross platform agreement and longitudinal consistency across tasks and system configurations. Energy based measures were more sensitive to microphone configuration, while perturbation and cepstral measures showed poor agreement across systems. These findings suggest that acoustic measures should not be treated as equally portable across platforms.

These results are broadly consistent with prior work. Amir et al. reported stronger cross-platform reliability for frequency than perturbation measures when comparing MDVP and Praat [8]. Kang et al. demonstrated Praat-based analysis as a functional alternative to CSL for select features [5]. Burris et al. and Smits et al. both documented that central tendency measures consistently outperform perturbation-based ones across systems [14,16]. The present study extends this literature by combining concurrent recordings across three consecutive days with a four-condition hardware and software analysis, allowing the contributions of microphone and analysis pipeline to be examined separately.

The disagreement observed across features pointed to several identifiable sources. For energy features, microphone related differences appeared to be the strongest driver. Agreement improved substantially when the same microphone signal was analyzed across software platforms, indicating that energy measures are highly sensitive to recording hardware and microphone placement. In the primary comparison, the headset microphone was positioned closer to the mouth and likely had a different frequency response and sensitivity profile than the CSL microphone, directly influencing absolute amplitude values [10,11,12]. Platform level differences in energy scaling may have added further variability. For pitch-based measures, differences in pitch floor and ceiling settings likely influenced the range within which pitch periods were detected. This may have contributed to the poor and inconsistent agreement seen in Pitch Maximum and Pitch Range across tasks and conditions. The stronger agreement observed during sustained vowel tasks is likely because sustained phonation produces a more stable and predictable vocal signal than connected speech. In connected speech, rapid changes in pitch and energy may be handled differently across platforms. Absolute Pitch Minimum and Pitch Maximum values may also be disproportionately influenced by brief tracking errors or outlier values. For this reason, percentile-based measures, such as the 95th percentile of pitch, may be more robust than absolute minimum and maximum values for connected speech tasks. These analysis parameters should be configured carefully and kept consistent throughout use. Perturbation measures, including jitter, shimmer, and CPP, showed consistently poor agreement across every task and configuration. This finding is consistent with prior literature showing that perturbation and cepstral measures are sensitive to differences in pitch period detection, signal processing algorithms, recording quality, and analysis settings [8,13,14,15,16]. CSL and Praat-Parselmouth use different algorithms for pitch period detection and even small differences in cycle boundary identification can produce meaningfully divergent perturbation values. These measures should therefore be interpreted within a consistent recording and analysis framework.

The main methodological limitations were manual trimming and differences in default analysis settings across systems. Manual segmentation may have introduced small differences in task boundaries, especially at onset and offset regions. Differences in pitch floor and ceiling settings may also have contributed to disagreement in Pitch Minimum, Pitch Maximum and Pitch Range.

These findings provide practical guidance for the next stage of LIS-N development. Future versions should incorporate automated trimming to improve segment consistency, standardized instructions for microphone distance, harmonized pitch settings and percentile-based pitch metrics for connected speech. As LIS-N moves toward full mobile deployment, these results can inform recording guidance and analysis settings for remote longitudinal voice monitoring. Future work would also evaluate usability and performance across more diverse populations, including different age groups and clinical conditions, to support broader generalizability. Clinical-grade assessment should continue to use consistent device and software ecosystems, especially when comparing acoustic values over time.

## 6. Conclusion

This study provides proof of concept for the viability of open-source acoustic analysis frameworks as accessible alternatives to proprietary clinical systems for selected acoustic measures. The LIS-N prototype demonstrated strong agreement with CSL for frequency-based measures and showed consistent longitudinal tracking across recording sessions. However, each platform operates within its own recording and signal processing framework, so values should be interpreted within the same system rather than mixed across systems. Microphone standardization and consistent analysis settings should be treated as protocol requirements in remote acoustic assessment. These findings support further development of Praat-Parselmouth within LIS N as a foundation for remote voice monitoring, including pre and post operative voice tracking and other telehealth applications.

## Conflict of Interest

The authors have no conflicts of interest to disclose

## Funding/Support Sources

The corresponding author is a salaried employee of the University of South Florida. This study and the app development form part of the work supported by Bridge2AI-Voice funding grant number – OT2-OD032720-01S3

## AI use disclosure

The authors used Claude Sonnet 4.5/4.6(Anthropic) [October 2025 – June 2026] for manuscript drafting and copy-editing assistance during the preparation of this manuscript. All scientific claims, analyses, interpretations, and final wording are the authors’ own, and the authors take full responsibility for the content of the manuscript. AI tools were also used to assist in designing the application. No AI tool was used to generate analysis code that produced reportable results without author review.

## Data Availability Statement

The data supporting the findings of this study are not publicly available due to participant privacy and confidentiality restrictions. Data access is limited to the approved study team and governed by institutional review requirements.

## Bridge2AI-Voice Consortium members

The following individuals are members of the Bridge2AI-Voice Consortium and contributed to the design, recruitment, data collection, ethical oversight, and infrastructure of the Bridge2AI-Voice study(named co-authors above are not re-listed here):

Alexandros Sigaras (Weill Cornell Medicine, New York, NY, USA); Anais Rameau (Weill Cornell Medicine, New York, NY, USA); Olivier Elemento (Weill Cornell Medicine, New York, NY, USA); Maria Powell (Vanderbilt University Medical Center, Nashville, TN, USA); David Dorr (Oregon Health & Science University, Portland, OR, USA); Phillip Payne (Washington University in St. Louis School of Medicine, St. Louis, MO, USA); Vardit Ravitsky (The Hastings Center, Garrison, NY, USA); Jean-Christophe Bélisle-Pipon (Simon Fraser University, Burnaby, BC, Canada); Ruth Bahr (University of South Florida, Tampa, FL, USA); Donald Bolser (University of Florida, Gainesville, FL); Jennifer Siu (Hospital for Sick Children, Toronto, ON, Canada); Jordan Lerner-Ellis (University of Toronto, Toronto, ON, Canada); Frank Rudzicz (Dalhousie University; Vector Institute, Halifax, NS, Canada); Micah Boyer (University of South Florida, Tampa, FL, USA); Yassmeen Abdel-Aty (University of South Florida, Tampa, FL, USA); Toufeeq Ahmed Syed (UTHealth Houston, Houston, TX, USA); James Anibal (NIH Clinical Center, U.S. National Institutes of Health; Institute of Biomedical Engineering, University of Oxford, Bethesda, Maryland, USA); Dona Amraei (Simon Fraser University, Burnaby, BC, Canada); Stephen Aradi (University of South Florida, Tampa, FL, USA); Kirollos Armosh (University of South Florida, Tampa, FL, USA); Ana Sophia Avila Martinez (University of South Florida, Tampa, FL, USA); Shaheen Awan (University of Central Florida, Orlando, FL, USA); Steven Bedrick (Oregon Health & Science University, Portland, OR, USA); Helena Beltran (University of South Florida, Tampa, FL, USA); Alexander Bernier (Simon Fraser University, Burnaby, BC, Canada); Moroni Berrios (University of South Florida, Tampa, FL, USA); Isaac Bevers (Massachusetts Institute of Technology, Cambridge, MA, USA); Alden Blatter (Simon Fraser University, Burnaby, BC, Canada); Rahul Brito (Harvard University; Massachusetts Institute of Technology, Boston, MA, USA); Amy Brown (Vanderbilt University Medical Center, Nashville, TN, USA); John Brown (University of South Florida, Tampa, FL, USA); Léo Cadillac (Simon Fraser University, Burnaby, BC, Canada); Selina Casalino (Mount Sinai Hospital, Sinai Health, Toronto, ON, Canada); Amanda Chao (Baycrest Centre, North York, ON, Canada); John Costello (Boston Children’s Hospital, Boston, MA, USA); Abhijeet Dalal (Oregon Health & Science University, Portland, OR, USA); Iris De Santiago (University of South Florida, Tampa, FL, USA); Enrique Diaz-Ocampo (CENIDET, Cuernavaca, Mexico); Amanda Doherty-Kirby (Simon Fraser University, Burnaby, BC, Canada); Ellie Eiseman (University of South Florida, Tampa, FL, USA); Mahmoud Elmahdy (University of South Florida, Tampa, FL, USA); Renee English (Simon Fraser University, Burnaby, BC, Canada); Emily Evangelista (University of South Florida, Tampa, FL, USA); Kenneth Fletcher (Vanderbilt University Medical Center, Nashville, TN, USA); Hortense Gallois (Simon Fraser University, Burnaby, BC, Canada); C. Gaelyn Garrett (Vanderbilt University Medical Center, Nashville, TN, USA); Alexander Gelbard (Vanderbilt University, Nashville, TN, USA); Omar Ghaffar (Hennick Bridgepoint Hospital, Toronto, ON, Canada); Amer Ghavanini (Trillium Health Partners and University of Toronto, Toronto, ON, Canada); Anna Goldenberg (The Hospital for Sick Children, Toronto, ON, Canada); Karim Hanna (University of South Florida, Tampa, FL, USA); William Hersh (Oregon Health & Science University, Portland, OR, USA); Jennifer Jain (University of South Florida, Tampa, FL, USA); Lochana Jayachandran (Sinai Health, Toronto, ON, Canada); Kaley Jenney It is made available under a CC-BY 4.0 International license. (which was not certified by peer review) is the author/funder, who has granted medRxiv a license to display the preprint in perpetuity. medRxiv preprint doi: https://doi.org/10.64898/2026.05.28.26354346; this version posted May 30, 2026. The copyright holder for this preprint (Massachusetts Institute of Technology, Cambridge, MA, USA); Kathy Jenkins (Boston Children’s Hospital, Boston, MA, USA); Stacy Jo (Boston Children’s Hospital, Boston, MA, USA); Alistair Johnson (University of Toronto, Toronto, ON, Canada); Brenda Juan Guardela (University of South Florida, Tampa, FL, USA); Ayush Kalia (University of South Florida, Tampa, FL, USA); Megha Kalia (University of South Florida, Tampa, FL, USA); Zoha Khawaja (Simon Fraser University, Burnaby, BC, Canada); Kenji Kobayashi (Vanderbilt University Medical Center, Nashville, TN, USA); Cynthia Kostelnik (University of South Florida, Tampa, FL, USA); Alisa Krause (University of South Florida, Tampa, FL, USA); Andrea Krussel (Washington University in St. Louis, St. Louis, MO, USA); Elisa Lapadula (Sinai Health, Toronto, ON, Canada); Genelle Leo (University of South Florida, Tampa, FL, USA); Justin Levinsky (Hospital for Sick Children, Toronto, ON, Canada); Chloe Loewith (Simon Fraser University, Burnaby, BC, Canada); Linda Ma (Baycrest Centre, North York, ON, Canada); Radhika Mahajan (Mount Sinai Hospital, Sinai Health, Toronto; Lunenfeld-Tanenbaum Research Institute, Sinai Health, Toronto, Toronto, ON, Canada); Vrishni Maharaj (University of South Florida, Tampa, FL, USA); Marie-Françoise Malo (Simon Fraser University, Burnaby, BC, Canada); Siyu Miao (The Hospital for Sick Children, Toronto, ON, Canada); LeAnn Michaels (Oregon Health & Science University, Portland, OR, USA); Matthew Mifsud (University of South Florida, Tampa, FL, USA); Marian Mikael (University of South Florida, Tampa, FL, USA); Pablo Montoya Varela (Simon Fraser University, Burnaby, BC, Canada); Elijah Moothedan (Florida Atlantic University, Boca Raton, FL, USA); Yosef Nafii (University of South Florida, Tampa, FL, USA); Tempestt Neal (University of South Florida, Tampa, FL, USA); Karlee Newberry (University of South Florida, Tampa, FL, USA); Evan Ng (The Hospital for Sick Children, Toronto, ON, Canada); Christopher Nickel (University of South Florida, Tampa, FL, USA); Amanda Peltier (Vanderbilt University Medical Center, Nashville, TN, USA); Trevor Pharr (University of South Florida, Tampa, FL, USA); Michaela Pnacekova (Simon Fraser University, Burnaby, BC, Canada); Matthew Pontell (Vanderbilt University Medical Center, Nashville, TN, USA); Jaiden Potter (Simon Fraser University, Burnaby, BC, Canada); Claire Premi-Bortolotto (Simon Fraser University, Burnaby, BC, Canada); Parnaz Rafatjou (University of South Florida, Tampa, FL, USA); JM Rahman (University of South Florida, Tampa, FL, USA); Gayathiri Rajkumar (Baycrest Centre, North York, ON, Canada); John Ramos (Weill Cornell Medicine, New York, NY, USA); Sarah Rohde (Vanderbilt University Medical Center, Nashville, TN, USA); Michael de Riesthal (Vanderbilt University Medical Center, Nashville, TN, USA); Jillian Rossi (University of South Florida, Tampa, FL, USA); Laurie Russell (Hospital for Sick Children, Toronto, ON, Canada); Samantha Salvi Cruz (Vanderbilt University Medical Center, Nashville, TN, USA); Joyce Samuel (Hospital for Sick Children, Toronto, ON, Canada); Suketu Shah (University of South Florida, Tampa, FL, USA); Ahmed Shawkat (University of South Florida, Tampa, FL, USA); Elizabeth Silberholz (Boston Children’s Hospital, Boston, MA, USA); John Stark (University of South Florida, Tampa, FL, USA); Lala Su (Hospital for Sick Children, Toronto, ON, Canada); Duncan Sutherland (New York, NY, USA); Venkata Swarna Mukhi Talluri (Oregon Health & Science University, Tempe, AZ, USA); Jeffrey Tang (Weill Cornell Medicine, New York, NY, USA); Luka Taylor (Simon Fraser University, Burnaby, BC, Canada); Jamie Toghranegar (University of South Florida, Tampa, FL, USA); Julie Tu (Mount Sinai Hospital, Toronto, ON, Canada); Megan Urbano (University of South Florida, Tampa, FL, USA); Gavin Victor (Simon Fraser University, Burnaby, BC, Canada); Kimberly Vinson (Vanderbilt University Medical Center, Nashville, TN, USA); Jordan Wilke (Massachusetts Institute of Technology, Cambridge, MA, USA); Claire It is made available under a CC-BY 4.0 International license. (which was not certified by peer review) is the author/funder, who has granted medRxiv a license to display the preprint in perpetuity. medRxiv preprint doi: https://doi.org/10.64898/2026.05.28.26354346; this version posted May 30, 2026. The copyright holder for this preprint Wilson (Simon Fraser University, Burnaby, BC, Canada); Madeleine Zanin (Mount Sinai Hospital, Toronto, ON, Canada); Xijie Zeng (Dalhousie University & Vector Institute, Halifax, NS, Canada); Theresa Zesiewicz (University of South Florida, Tampa, FL, USA); Robin Zhao (Weill Cornell Medicine, New York, NY, USA); Pantelis Zisimopoulos (Weill Cornell Medicine, New York, NY, USA); Satrajit Ghosh (Massachusetts Institute of Technology, Cambridge, MA, USA).

## Notes

### Competing Interest Statement

The authors have declared no competing interest.

### Author Declarations

Institutional Review Board of the University of South Florida deemed this study exempt and Not Human Research Subject

